# Investigations on Clinical Validation of mutated ANKRD36 as a Novel Molecular Biomarker for Monitoring Early Progression and Timely Therapeutic Interventions in Blast Crisis CML

**DOI:** 10.1101/2023.12.28.23300606

**Authors:** Nawaf Alanazi, Abdulaziz Siyal, Muhammad Absar, Masood Shammas, Amer Mahmood, Sulman Basit, Sarah AlMukhaylid, Zafar Iqbal

## Abstract

**Background:** Chronic Myeloid Leukemia (CML) is initiated in the bone marrow due to the chromosomal translocation t(9;22), resulting in the fusion oncogene BCR-ABL. Tyrosine kinase inhibitors (TKIs) targeting BCR-ABL have transformed fatal CML into an almost curable disease. However, TKIs lose efficacy during disease progression, and the mechanism of CML progression remains to be fully understood. Additionally, common molecular biomarkers for CML progression are lacking. Our studies previously detected ANKRD36 (c.1183_1184 delGC and c.1187_1188 dupTT) associated exclusively with advanced phase CML. However, clinical validation of this finding was pending. Therefore, this study aimed to clinically validate mutated ANKRD36 as a novel biomarker of CML progression.

**Materials and Methods:** The study enrolled 124 patients in all phases of CML, recruited from Mayo Hospital and Hameed Latif Hospital in Lahore, Punjab, between January 2020 and September 2023. All response criteria were adopted from the European LeukemiaNet guideline 2020. Informed consent was obtained from all study subjects. The study was approved by scientific and ethical review committees of all participating centers.

Sanger sequencing was employed to detect ANKRD36 mutations in CML patients in accelerated phase (AP) (n=11) and blast crisis (BC) (n=10), with chronic-phase CML (CP-CML) patients as controls (n=103). Samples were processed using Big Dye Terminator Cycle Sequencing Ready Reaction kits and sequenced using ABI Prism 3730 Genetic Analyzer, and sequencing using forward and reverse primers for ANKRD36. Data was analyzed using SPSS version 26.

**Results:** During our study, 17% of CML patients progressed to advanced phases AP-CML n= 11 (8.9%) and BC-CML n=10 (8.1%). The chronic- and advanced-phase patients showed significant difference with respect to male-to-female ratio, hemoglobin level, WBC count, and platelet count. Sanger sequencing detected ANKRD36 mutations c.1183_1184 delGC and c.1187_1185 dupTT exclusively in all AP- and BC-CML patients but in none of the CP-CML patients. Nevertheless, mutations status was not associated with male-to-female ratio, hemoglobin level, WBC count, and platelet count, which makes ANKRD32 as an independent predictor of early and terminal disease progression in CML.

**Conclusions:** The study confirms ANKRD36 as a novel genomic biomarker for early and late CML progression. Further prospective studies should be carried out in this regard. ANKRD36, although fully uncharacterized in humans, shows the highest expression in bone marrow, particularly myeloid cells and correlation with high WBC count. Functional integrated genomic studies are recommended to further explore the role of ANKRD36 in the biology and pathogenesis of CML.

## 1. Introduction

Chronic myeloid leukemia (CML) is a clonal myeloproliferative neoplasm of the hematopoietic stem cells (HSCs), marked by the uncontrolled excessive production of dysfunctional granulocytes and progenitors’ cells, in the bone marrow and peripheral blood (1). CML is the first aberrant chromosome abnormality described in a malignancy, known as the Philadelphia (Ph+) chromosome (2). The presence of Philadelphia chromosome t(9:22), which is a chromosomal reciprocal translocation between Abelson murine leukaemia (ABL) on the long arm of chromosome 9 and breakpoint cluster region (BCR) on chromosome 22, is a defining feature of CML (2,3). As a result of BCR-ABL1 fusion oncoprotein (P210) and the constitutive activity of tyrosine kinase, hematopoietic stem cells (HSC) are expanded clonally and CML is induced (4,5).

CML is a triphasic course disease classified into Chronic Phase (CP), Accelerated Phase (AP), and Blast Crisis (BC) (6). In the chronic phase (CP-CML) of chronic myeloid leukemia, leukemia cells can mature into granulocytes. However, as the disease advances, it transitions to the accelerated phase (AP-CML) and rapidly progresses to blast crisis (BC-CML). During blast crisis, there is a surge in aggressive and immature blast cells, leading to a loss of the ability to differentiate cells (7). Unfortunately, despite all advancements in CML treatment, from 1st generation TKIs to Asciminib in the last two decades, the outcome of BC-CML is dismal, and overall survival is few months only (8,11,12). BC-CML is a result of genetic instability and mutation acquisition sparked by the over-expression of BCR-ABL1 and resistance to tyrosine kinase inhibitors (TKIs) thus causing drug resistance and leading to morbidities and mortality (9,10). Due to genetic instability, BC-CML is marked by the presence of additional chromosomal aberrations leading to notable restrictions in the effectiveness of TKIs for treating BC-CML (11). Therefore, there is a pressing need for the identification and characterization of more precisely defined molecular biomarkers. This is crucial for the development of improved treatment modalities tailored to the specific characteristics of BP-CML, aiming to enhance therapeutic outcomes.

Furthermore, the lack of biomarkers for CML progression is attributed to a limited comprehension of the underlying mechanisms. In our previous studies on advanced CML patients, we identified ankyrin repeat domain 36 (ANKRD36) gene as a potential, novel and common biomarker of disease progression in CML patients and hence it may serve as a biomarker to identify CML patients at risk of progression (13). Nevertheless, this finding needed clinical validation. Therefore, the objective of this study was to clinically validate previously detected ANAKRD36 mutation as a novel biomarker to monitor CML progression and timely intervene therapeutically.

## 2. Materials and Methods

### 2.1. Patient Inclusion and Exclusion Criteria

The study was carried out among Chronic Myeloid Leukemia (CML) patients recruited from two medical centers Mayo Hospital and Hameed Latif Hospital in Lahore, Punjab from January 2020 to September 2023. The number of CML patients included in this study was 124. The experimental group comprised 11 Accelerated Phase (AP-CML) patients and 10 BC-CML while 103 age/gender-matched Chronic Phase (CP-CML) patients were used as controls. All CP-CML patients received imatinib mesylate as first-line treatment and those who exhibited resistance to IM were subsequently administered nilotinib (NI), as dasatinib and ponatinib were not available. Treatment response criteria were determined according to the European Leukemia Net guidelines 2020 (14).

### 2.2. Ethical Approval

The study adhered to the principles outlined in the Declaration of Helsinki. Written informed consent was obtained from all participating patients. The approval of study protocols was obtained from Scientific Committees and Ethical Review Boards (ERBs) of King Abdullah International Medical Research Center (KAIMRC); King Saud bin Abdulaziz University for Health Sciences (KSAU-HS), Hayatabad Medical Complex (HMC), Mayo Hospital, and Hameed Latif Hospital in Lahore, Punjab, Pakistan.

### 2.3. Sample Collection and DNA Extraction

Ten milliliters of peripheral blood samples were collected in 3-5ml EDTA tubes (BD Vacutainer Systems, Franklin Lakes, N.J.) from all age groups and clinical phases of CML biweekly, for Follow-up and medication refills, during their visits to the outpatient department (OPD) of the medical oncology unit, Mayo Hospital, and Hameed Latif Hospital in Lahore, Punjab. DNA extraction from all patients was carried out using the QIAamp DNA Blood Mini Kit (QIAGEN). The quantification of DNA was conducted using the NanoDrop Spectrophotometer (NanoDrop Technologies, Inc., Wilmington, DE, USA). Subsequently, DNA was diluted into aliquots of 70– 80 ng/μL for the detection of mutations. The surplus DNA was further diluted to 40 ng/μL for Sanger sequencing. Storage of DNA was performed in a freezer at −80 °C (13).

### 2.4. Mutation detection by Sanger Sequencing

Amplification of samples was done by PCR using forward and reverse primers, as reported previously (13). Sequencing reactions were prepared using ABI PRISM Big Dye Terminator Cycle Sequencing Ready Reaction kits. The sequencing was carried out using ABI Prism 3730 Genetic Analyzer (Applied Biosystems, California, USA).

### 2.5. Statistical Analysis of Patient Clinical Data

Categorical variables were represented with percentages and absolute numbers, while continuous variables were measured with mean and median according to the normality test. Chi-Square and Fisher’s exact test were utilized to compare the categorical data of two groups, depending on applicability, while the comparison of two groups of continuous data was done by two-sample independent test or Mann Whitney U test.

## 3. Results

### 3.1. Demographic and Clinical Description of Patients

This study encompassed a cohort of 124 Chronic Myeloid Leukemia (CML) patients, with an average patient age of 35.9 years (Table 1). The gender distribution showed a male-to-female ratio of 1:1. Further analysis of gender demographics disclosed that females constituted (n=59) 47.5% of the patients, while males accounted (n=65) for 52.4%. The average hemoglobin level was 11.3, and the mean White Blood Cell (WBC) count was 152.27. Additionally, the platelet count for CML patients was measured at 358.72. In summary, the prevalence of CML was higher among males based on these observations.

**Table 1.**
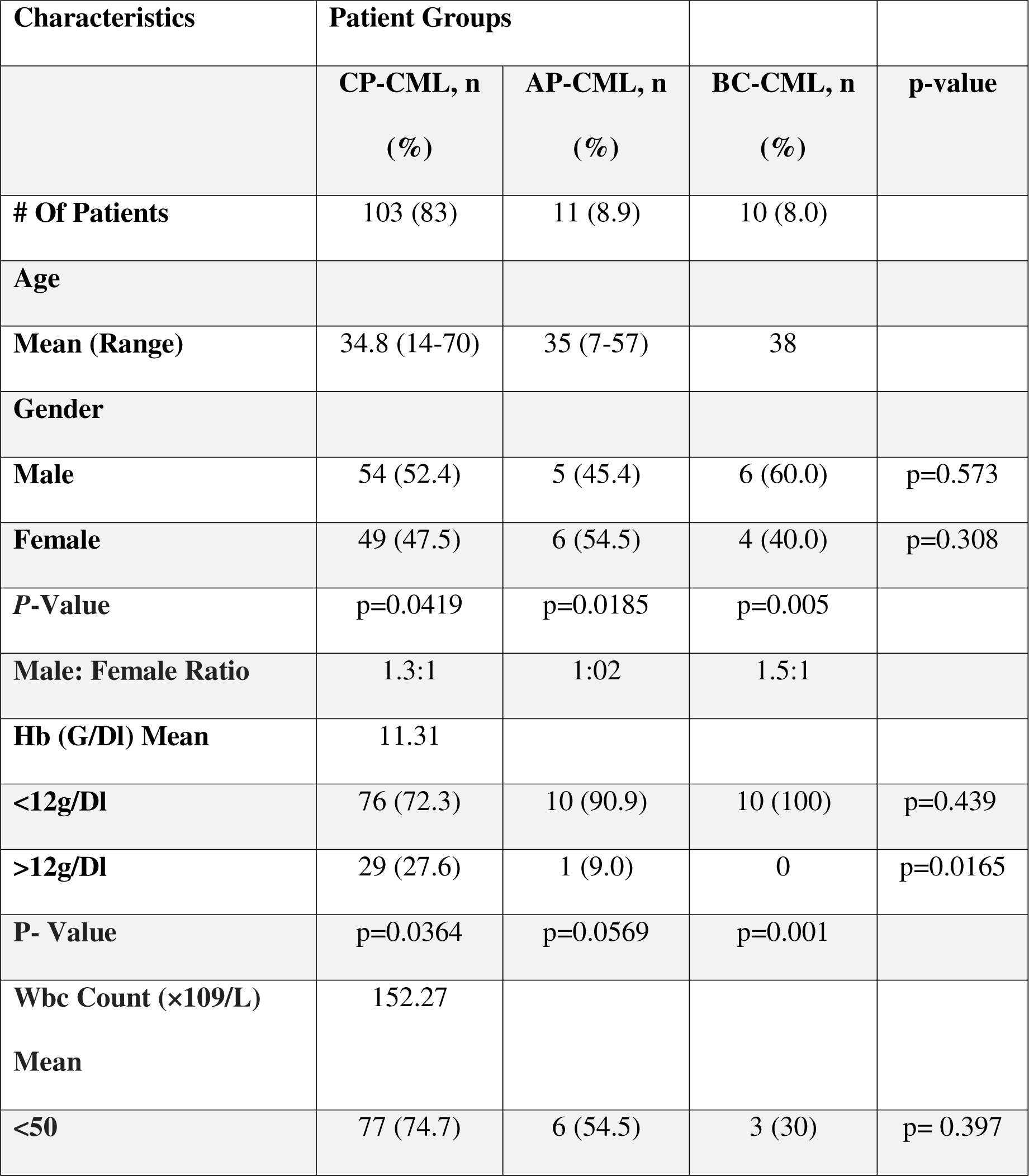
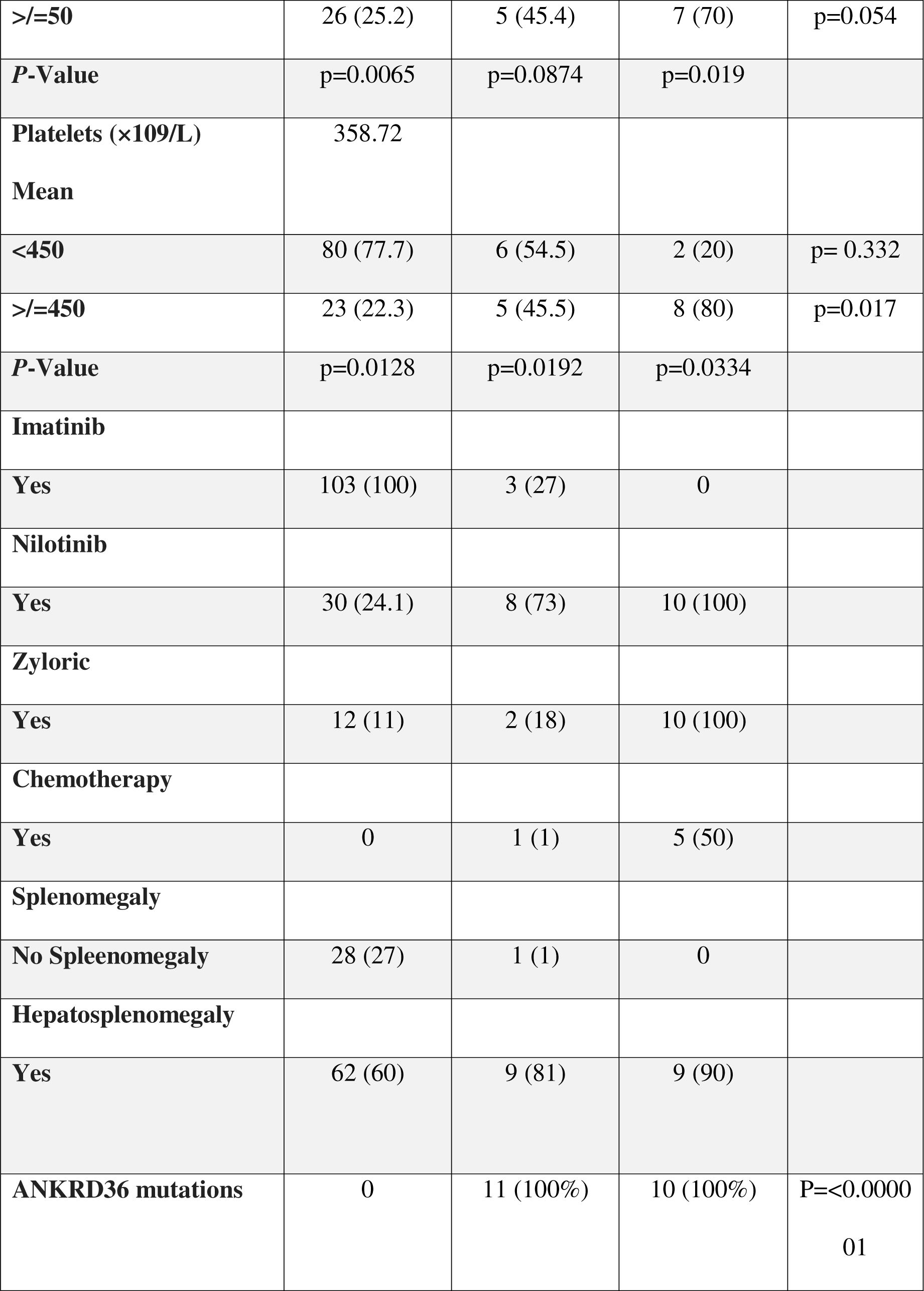
Comparison of demographics, clinical data, and laboratory parameters between three phases of CML.

During the study, 17% (n = 21) of patients progressed to advanced phases out of which AP-CML comprised 11 (8.9%) and BC 10 (8.1%). CP-, AP- and BC-CML patients had mean ages of 34.8, 35.0, and 38.0 years, respectively. Notably, male dominance was observed in all phases of CML except for AP-CML. The calculated male-to-female ratios were 1.3:1, 1:02, and 1.5:1 in BC, AP, and CP, respectively. Additionally, anemia was prevalent among three-fourths of the patients. Approximately 30.6% of all CML patients exhibited a leukocyte count of 50 × 10^9/L or higher (n = 38). Imatinib served as the first-line Tyrosine Kinase Inhibitor (TKI), administered to 100% and 27% of CP and AP patients, respectively. In cases of TKI resistance and/or progression to blast crisis, nilotinib was drug of choice, with usage percentages in CP-CML (24.1%), AP-CML (73%), and BC-CML (100) %. Overall, 17% of CML patients (n = 21) progressed to either AP-CML (n = 11) or BC-CML (n = 10) (refer to Table 1).

There was a significant difference between chronic- and advanced-phase patients concerning male-to-female ratio, hemoglobin level, WBC count, and platelet count (Table 1).

### 3.2. Mutation detection by Sanger Sequencing

Sanger sequencing confirmed the presence of ANKRD36 gene mutations, (c.1183_1184 delGC and c.1187_1188 dupTT) (Figure 1), in patients with BC-CML (n= 10), indicating a correlation between ANKRD36 variants and the progression of Chronic Myeloid Leukemia (CML). Additionally, the confirmation of ANKRD36 mutations in Accelerated Phase CML (AP-CML) which was found in all AP-CML progressed patients (n=11), suggests that these mutations serve as an early indication of CML progression, highlighting their potential as early biomarkers for disease advancement. This mutation was not detected in any of the CP-CML patients who served as controls (n=103), which shows that it is exclusively associated with AP-/BC-CML. Moreover, mutational status was not associated with any of the demographic and clinical parameters which makes mutated ANKRD32 an independent predictor of early and terminal disease progression in CML.

**Figure 1.**
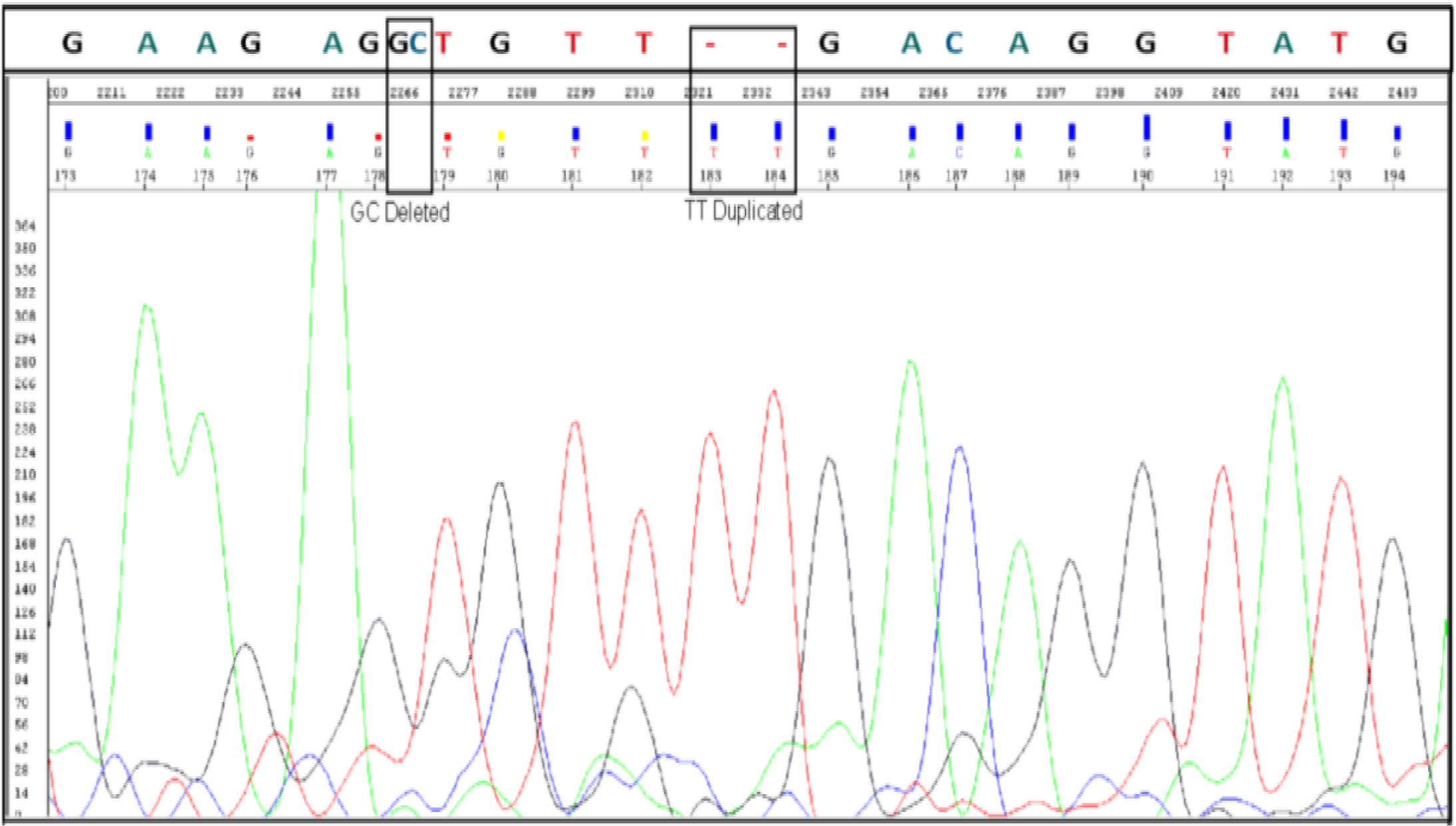
Confirmation of c.1183_1184 delGC and c.1187_1185 dup TT mutation in ANKRD36 gene by Sanger sequencing in Accelerate/Blast phase (AP/BC) CML patients (AP, n=11, BC, n=10).

Therefore, it is concluded that ANKRD36 is a novel molecular biomarker for monitoring accelerated phase (early progression) and blast crisis (terminal progression) in CML patients.

## 4. Discussion

The progression of Chronic Myeloid Leukemia (CML) into advanced phases (-AP and -BC), results in drug resistance, treatment failure, comorbidities, and increased mortality (9). Identifying genetic variants associated with advanced phases becomes crucial for early detection of fatal disease progression, enabling timely clinical intervention to delay or prevent acute transformation in CML. As we have reported ANKRD36 as a novel gene associated with CML progression (13). To clinically validate the potential of ANKRD36 as a novel biomarker of CML progression, eligible CML patients in advanced phases, i.e., accelerated and blast crisis (AP-/BC-CML), were subjected to Sanger sequencing alongside CP-CML controls in the current study.

This study included 124 patients from all three phases of CML. In our study, the mean age of the patients was 35.9 years. The mean age of our CML patients significantly differs from that of Western populations. In Europe, the mean age of CML patients was reported to be 57-60 years, while a study from the USA indicated a mean age of 56 years (15,16). In Japan, the mean age of patients diagnosed with CML was 56 years (17). In our study, females constituted 47.5% (n=59) of the patients, while males accounted for 52.4% (n=65).

Imatinib was the first-line treatment for 100% (n=103) and 27% (n=3) of patients in the Chronic Phase (CP) and Accelerated Phase (AP), respectively. In TKI-resistant cases and patients who progressed, nilotinib was the primary choice, used in 24.1% (n=30) for CP-CML, 73% (n=8) for AP-CML, and 100% (n=10) for BC-CML. Similarly, in a French population study involving 3,633 patients, imatinib emerged as the primary first-line treatment, administered to 77.6% of individuals (n = 2,821), followed by nilotinib for 18.3% (n = 663) (18). Overall, 17% of CML patients (n = 21) progressed to either AP-CML (n = 11) or BC-CML (n = 10). A study conducted and all approved TKIs in technologically advanced countries (2,3,13). Furthermore, there was a significant difference between chronic- and advanced-phase patients concerning male-to-female ratio, hemoglobin level, WBC count, and platelet count. Nevertheless, these parametric patients at risk of developing disease progression (8, 11).

In our previous report, we discovered a novel biomarker associated with the progression of Chronic Myeloid Leukemia (CML) (13). The current study focuses on the clinical validation of progression. This could be due to very important biological role of this gene (20–26).

ANKRD36 belongs to a group of genes called Ankyrin repeat domain-containing proteins (ANKRD) (20). ANKRD genes are widely observed in humans, with around 270 known proteins containing these common protein-protein interaction domains, each serving various functions (20). Positioned on chromosome 2, ANKRD36 is characterized by 36 exons and encompasses six repeating units (20). These units are composed of two antiparallel helices and a hairpin structure, forming a repetitive stacking pattern on the superhelix (20). Earlier, our protein bio-modeling studies indicate that ANKRD36 mutation (c.1183_1184 delGC and c.1187_1188 dupTT). The simultaneous “deletion of GC and insertion of TT” results in two amino acid changes: Ala to Cys (395) and Val to Phe (396). Both Val and Phe, being hydrophobic and positionally interchangeable, maintain the same overall protein function due to the retention of specific nucleotides in the DNA codon, encoding amino acids with similar polarity or hydrophobicity substitution (13). However, the A395C mutation has not been previously reported, emphasizing its potential significance as rare mutations often exhibit higher pathogenicity than more common ones. The mutation is located on the surface exterior, linking two alpha helices, and may alter the flexibility of the protein, potentially impacting interactions with other proteins (13). Nevertheless, in-depth investigations into the functions of ANKRD36 are scarce, contributing to a lack of understanding regarding its involvement in human diseases. However, ANKRD36 has been reported to be associated with various health conditions and malignancies.

A study reported by Jiang et al. on Myocardial Infarction (MI) identified ANKRD36 as one of the five gene hubs expressed in MI patients, suggesting a potential association with various subtypes of MI (21). In patients with type 2 diabetes mellitus (T2DM), ANKRD36 expression was notably upregulated compared to normal controls. Expression levels of circANKRD36 were found to be increased in peripheral blood leucocytes, and this upregulation exhibited a correlation with chronic inflammation in T2DM patients, indicating its potential utility as a biomarker for screening chronic inflammation in T2DM (22). In hypertension, ANKRD36 is implicated in the regulation of blood pressure through its interaction with Y ing Yang 1 (YY1), consequently leading to alterations in the expression of ENaC genes (23). Moreover, ANKRD36 appears to play a potential role in the progression of Diabetic Kidney Disease (DKD) by influencing lipid metabolism and contributing to inflammation, as its expression is positively correlated with lipid profile and white blood cells (20).

By utilizing the online searching tool on “The Cancer Genome Atlas (TCGA)” and utilizing “cBioPortal for Cancer Genomics” from the National Cancer Institute of the National Institute of Health (Bethesda, MD, USA), we were unable to identify any leukemia-specific mutations in ANKRD36. Nonetheless, various studies have reported the role of ANKRD36 in different types of cancers. A study investigating the antitumor functions of miR-144-5p in renal cell carcinoma (RCC), indicated that miR-144-5p directly targets the ANKRD36 gene. The elevated expression of ANKRD36, regulated by miR-144-5p, was linked to poor survival in this study (24). Another study demonstrated that suppressing miR-182 led to an increase in apoptosis (25). Furthermore, the in-vivo experiment demonstrated a decrease in tumor growth when cells treated with anti-miR-182 were transplanted into immunodeficient mice (25). Furthermore, a correlation was identified between the mutational status of ANKRD36 genes and proximal gastric cancer (26). ANKRD36 has been reported to co-express and interact with other genes on locus 2q11.2, including ANKRD36C, ITPRIPL1, FAHD2B, FAM178B, and CNNM3 (27). Moreover, ANKRC36 has been associated with pro-inflammatory phenotype of fibroblast-like synoviocytes in Rheumatoid arthritis (28). This suggests that ANKRD36 is actively engaged in crucial biological networks associated with cancer.

Although further functional genomic studies are required to find out exact role of ANKRd36 mutations in CML progression, some recent reports shed light on its important role. According to the information obtained from the transcriptome analysis of the TCGA database, the expression of resting CD4 memory T cells is significantly elevated in patients with the ANKRD36 mutation (p = 0.032). Based on this finding, the authors of the study proposed that it is possible that the immune system of the patients with ANKRD36 mutations did not activate the immunological mechanism that is responsible for fighting tumors (29). We think that this could be one of the possible mechanisms through which ANKRD36 mutation might lead to CML progression.

All the above-mentioned findings highlight the importance of ANKRD36 in pivotal biological functions and its notable association with cancer progression in CML patients. Moreover, the identification of ANKRd36 mutations exclusively with AP-/BC-CML suggests that it is a novel molecular biomarker that can be used to monitor CML progression, early determination of patients at risk of CML progression, particularly blast crisis, and carry out timely therapeutic interventions of at-risk patients to delay or minimize blast crisis development, that can lead to considerable improvement in overall survival of high risk BC-CML patients (30). Given its highest expression in myeloid cells of the bone marrow, ANKRD36 holds potential as a potential novel drug target, particularly for patients with advanced phases of CML (13,31).

## 5. Conclusions

In conclusion, our studies show that CML progression is still a serious clinical issue, specifically in low-resourced countries. The challenge is related to early detection of patients at risk of developing BC-CML as well as timely therapeutic interventions, and could at least be partially addressed by discovery of novel molecular biomarkers of early determination of CML patients at risk of blast crisis. We showed that ANKRD36 gene mutations, c.1183_1184 delGC and c.1187_1188 dupTT), are exclusively associated with advanced phases of CML (AP-/ BC-CML) and therefore are very important biomarkers of early disease progression and blast crisis development in CML patients. Our prior protein biomodeling investigations indicate that these mutations alter the structure of the ANKRD36 protein, potentially influencing its biological functions. Despite the lack of comprehensive characterization in humans, various studies suggest the gene’s involvement in diverse biological functions and disease pathogenesis, including cancer. Given its highest expression in bone marrow, particularly in myeloid cells, and its correlation with high white blood cells count, the gene may have a significant role in hematopoiesis and potentially contribute to hematopoietic diseases if mutated, specifically in CML progression. Therefore, we advocate for further research to elucidate the precise biological functions of this gene, with a specific focus on its involvement in apoptosis and carcinogenesis in advanced phase CML.

## 6. Limitations

Validation of the identified biomarker was conducted in a limited number of patients, primarily due to the limited availability of AP/BC-CML patients. This scarcity could be attributed to the impact of the approach to six FDA-approved Tyrosine Kinase Inhibitors (TKIs), which has resulted in a reduction in the number of advanced-phase CML cases. Consequently, it is strongly recommended to undertake further validation of this significant biomarker in more extensive patient cohorts.

## Data Availability

All data produced in the present study are available upon reasonable request to the authors

## 7. Acknowledgments

The study was approved by King Abdullah International Medical Research Centre (KAIMRC), National Guard Health Affairs, Saudi Arabia, although no research funding was provided (project # RA17/002/A). This study was partially supported by the College of Medicine, Research Centre, Deanship of Scientific Research, King Saud University, Riyadh, Saudi Arabia. All authors have read and have consented to the acknowledgment.

